# Early Joint Trajectories of Liver-Related Laboratory Biomarkers and 60-Day Mortality in Sepsis-Associated Liver Injury

**DOI:** 10.64898/2026.06.27.26356734

**Authors:** Yanan Qi, Tian Zhou, Qinmin Zhao, Cheng Liu

## Abstract

**Background:** Sepsis-associated liver injury (SALI) is commonly assessed using static laboratory values, although liver dysfunction during sepsis is dynamic.

**Methods:** This retrospective cohort study included 162 ICU patients with SALI. Early trajectories of alanine aminotransferase, total bilirubin, and albumin during the first 7 days after ICU admission were identified using group-based multi-trajectory modeling. Landmark analysis and Cox regression were used to evaluate 60-day mortality.

**Results:** Twenty-five patients died within 60 days. Four trajectory classes were identified. Between-class separation was driven mainly by alanine aminotransferase and total bilirubin, whereas albumin showed limited short-term variation. After the landmark time point, Class 3 (HR, 4.374; 95% CI, 1.960-9.759; *P* <0 .001) and Class 4 (HR, 7.451; 95% CI, 3.649-15.212; *P* <0 .001) had higher mortality risk than Class 1.

**Conclusions:** Early joint trajectories of liver-related laboratory biomarkers may identify clinically meaningful SALI subphenotypes and support risk stratification in critically ill patients.

## Introduction

Sepsis is a life-threatening syndrome characterized by a dysregulated host response to infection, leading to acute organ dysfunction ^[1-3]^. Among the affected organs, the liver plays a central role in metabolic regulation, immune modulation, host defense, and drug/xenobiotic clearance. Sepsis-associated liver injury (SALI) is a common complication in critically ill patients and has been consistently associated with increased disease severity and poor clinical outcomes ^[4-8]^. Current approaches to the assessment of SALI primarily rely on static measurements of liver-related biomarkers, such as total bilirubin, alanine aminotransferase (ALT), and the international normalized ratio (INR) ^[6,9-11]^. While these markers are widely used in clinical practice, they are typically evaluated at single time points and may not adequately reflect the dynamic and heterogeneous nature of liver dysfunction during sepsis ^[8,12]^. The pathophysiology of SALI is complex and involves multiple processes, including cholestasis, hepatocellular injury, impaired synthetic function, and alterations in hepatic microcirculation ^[6-8,11,13]^. As a result, patients with similar baseline laboratory values may experience markedly different clinical trajectories ^[14]^. In recent years, trajectory-based analytical approaches have been increasingly applied in clinical and critical care research to characterize temporal patterns of physiological variables ^[15-18]^. Studies examining dynamic changes in lactate levels or Sequential Organ Failure Assessment (SOFA) scores have demonstrated that longitudinal patterns may provide additional prognostic information beyond single measurements ^[19]^. However, the application of such approaches to liver-related biomarkers in SALI remains limited. In particular, few studies have jointly modeled multiple dimensions of liver function to capture their combined dynamic evolution ^[16,20]^. Moreover, there is ongoing debate regarding whether early abnormalities in liver function or their subsequent progression are more relevant for predicting outcomes in sepsis ^[9-10,21]^. Some studies emphasize the prognostic value of peak or baseline bilirubin levels, whereas others suggest that persistent or worsening dysfunction over time may be more informative ^[9-11,22]^. These differing perspectives highlight the need for approaches that incorporate both baseline status and temporal changes. Therefore, the present study aimed to identify early joint trajectory patterns of ALT, total bilirubin, and albumin in patients with SALI using group-based multi-trajectory modeling ^[15-18,23]^. We further examined the association between these trajectory patterns and 60-day mortality ^[24-28]^. We hypothesized that distinct trajectory patterns exist and that patterns characterized by persistently elevated or progressively increasing bilirubin levels are associated with worse survival outcomes. By integrating longitudinal biomarker data, this study seeks to provide additional insights into the dynamic heterogeneity of SALI and its relationship with clinical outcomes.

## Materials and Methods

### Study Design and Population

This retrospective cohort study included patients admitted to the intensive care unit (ICU) at our institution between 2022-06-01 and 2024-12-30. Sepsis was defined according to the Sepsis-3 criteria. Sepsis-associated liver injury (SALI) was defined as total bilirubin >2 mg/dL in combination with an international normalized ratio (INR) >1.5 at any time during hospitalization ^[3,11]^. Patients were excluded if they were younger than 18 years, had pre-existing chronic liver disease, or had fewer than three repeated measurements of alanine aminotransferase (ALT), total bilirubin, or albumin within the first 7 days after ICU admission.

### Ethics Statement

The studies involving human participants were reviewed and approved by the Medical Ethics Committee of The First Affiliated Hospital of Bengbu Medical University (Approval No. [2023]342). The study was conducted in accordance with the principles of the Declaration of Helsinki. Written informed consent was not required because this was a retrospective study using anonymized clinical data collected during routine care. The data were fully anonymized prior to the researchers’ access, and no direct patient identifiers were obtained. The study did not involve direct patient contact, intervention, or collection of additional biological samples. The requirement for written informed consent was waived by the Medical Ethics Committee of The First Affiliated Hospital of Bengbu Medical University in accordance with national legislation and institutional requirements..

### Data Collection and Variables

Longitudinal laboratory measurements of ALT, total bilirubin, and albumin within the first 7 days after ICU admission were extracted from the electronic medical record system. To standardize time alignment across patients, laboratory data were organized into 48-hour time windows, with ICU admission defined as time zero. Three consecutive time points were selected for trajectory modeling based on data completeness. Baseline variables included demographic characteristics, liver-related laboratory parameters, coagulation indices, renal function markers, severity scores (including SOFA), organ support therapies (e. g., mechanical ventilation, CRRT), and comorbidities. The primary outcome was 60-day all-cause mortality, defined as death occurring within 60 days after ICU admission.

### Group-Based Multi-Trajectory Modeling

Group-based multi-trajectory modeling was used to identify distinct joint longitudinal patterns of ALT, total bilirubin, and albumin. The analysis was implemented in R using the gbmt package for group-based multivariate trajectory modeling. Models with two to five trajectory classes were fitted, and quadratic polynomial terms were used to describe temporal trends. Model selection was based on a combination of Bayesian information criterion (BIC), Akaike information criterion (AIC), sample size-adjusted BIC, average posterior probability (AvePP), odds of correct classification (OCC), minimum class size, and clinical interpretability. Each patient was assigned to the trajectory class with the highest posterior probability ^[15-18]^.

### Statistical Analysis

Continuous variables are presented as median (interquartile range), and categorical variables are presented as counts (percentages). Comparisons between groups were performed using the Kruskal-Wallis test for continuous variables and the chi-square test or Fisher’s exact test for categorical variables, as appropriate. Patients with fewer than three repeated measurements of ALT, total bilirubin, or albumin during the first 7 days were excluded before trajectory modeling. For regression analyses, variables with available data were used in complete-case models. Kaplan-Meier survival curves and landmark analysis were used to compare 60-day survival among trajectory classes ^[27]^. The day-22 landmark was selected as an exploratory time point based on the observed separation of survival curves and was used to reduce time-related bias when comparing post-landmark mortality. Cox proportional hazards regression models were used to evaluate the association between trajectory class membership and 60-day mortality ^[25]^. Multivariable models were adjusted for clinically relevant covariates selected a priori based on clinical experience, including age, lactate, renal disease, and SOFA score. The proportional hazards assumption was assessed using Schoenfeld residuals and weighted residual-based diagnostics ^[26,28]^. A two-sided P value <0.05 was considered statistically significant. All analyses were performed using R software (version 4.5.1).

## Results

### Baseline Characteristics

A total of 162 patients with sepsis-associated liver injury were included in the analysis, of whom 25 (15.4%) died within 60 days. The median age of the cohort was approximately 58 years, and 60.8% were male. Comparisons between survivors and non-survivors showed no significant differences in most baseline variables, including liver function indices, coagulation parameters, renal function, and organ support therapies. Albumin was the only liver-related biomarker that differed significantly between groups, with slightly higher levels observed in non-survivors (*P* = 0.037).

**Table 1.**
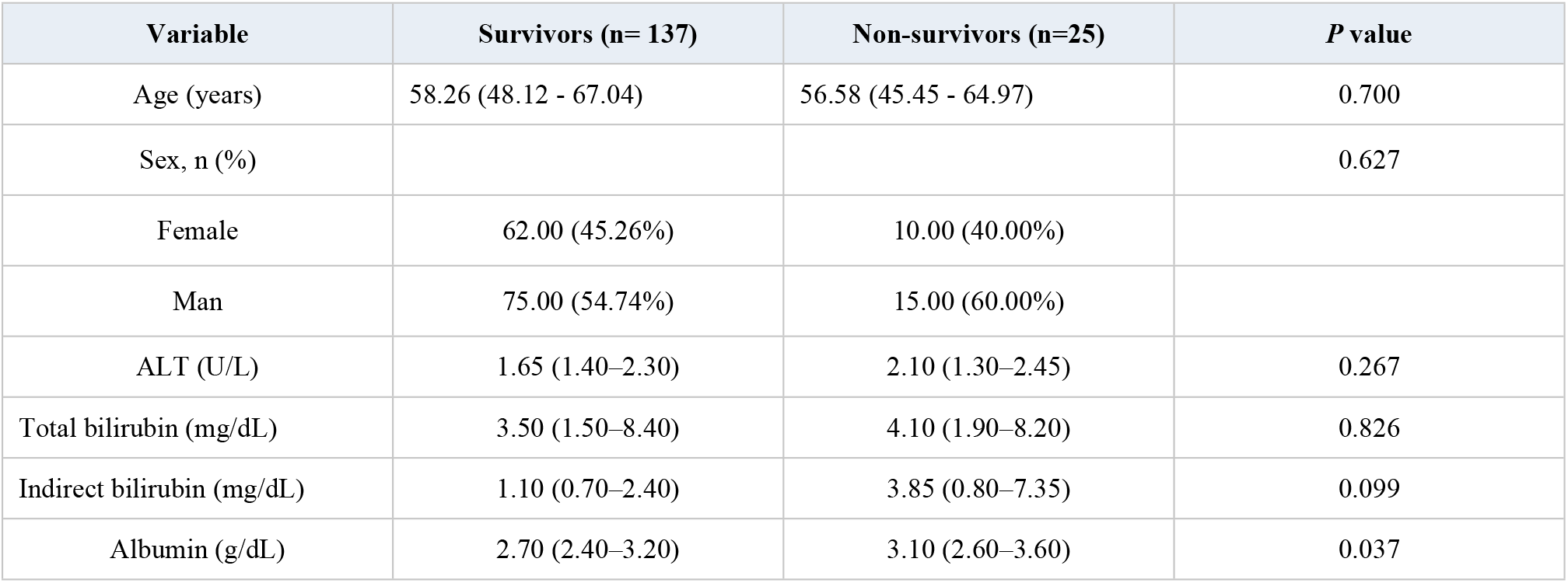
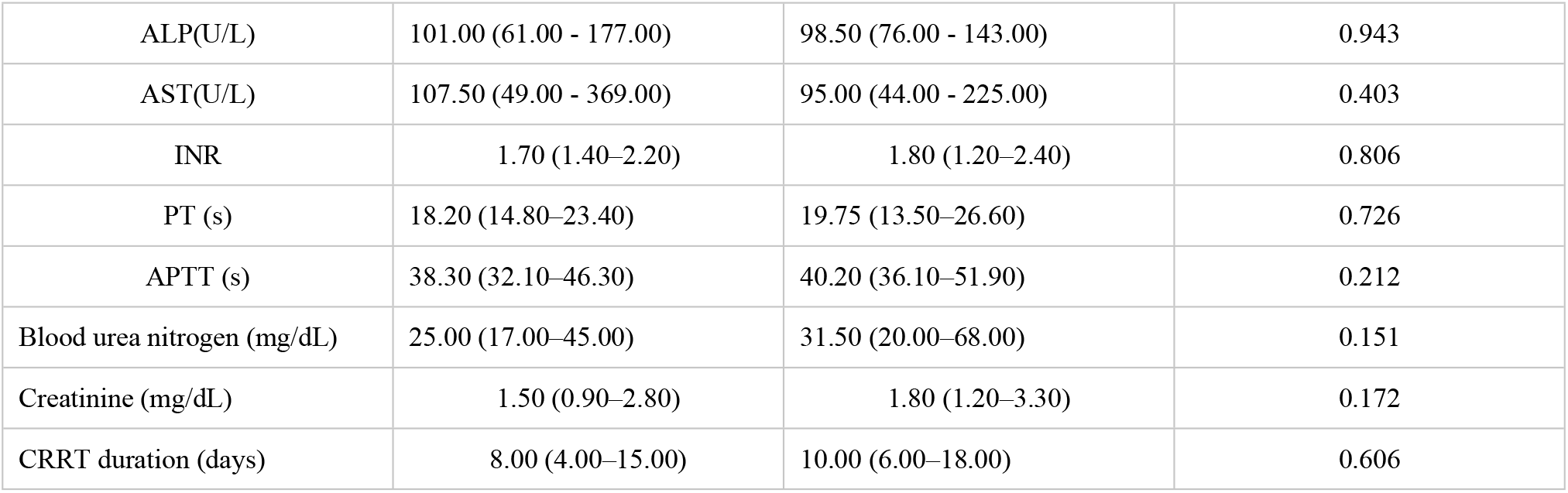
Baseline characteristics of the study participants according to 60-day survival status.

### Identification of Trajectory Classes

Group-based multi-trajectory modeling identified four distinct joint trajectory classes based on ALT, total bilirubin, and albumin. The proportions of patients in each group were 38.9% (Class 1), 15.4% (Class 2), 15.4% (Class 3), and 30.2% (Class 4). Model selection was based on multiple criteria, including BIC, AIC, average posterior probability, odds of correct classification, and clinical interpretability. The selected four-group model demonstrated acceptable classification performance across all trajectory classes.

**Table 2.**
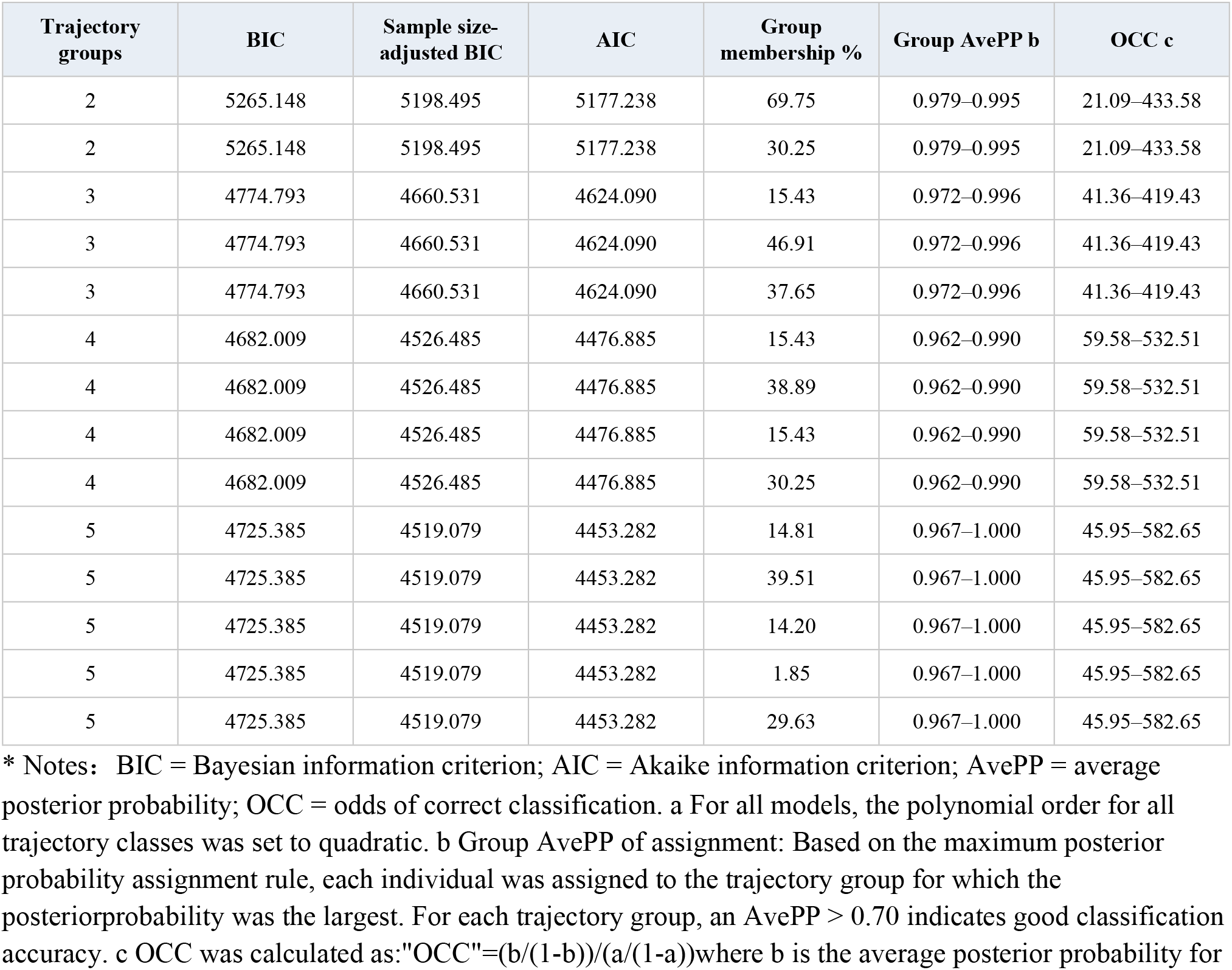
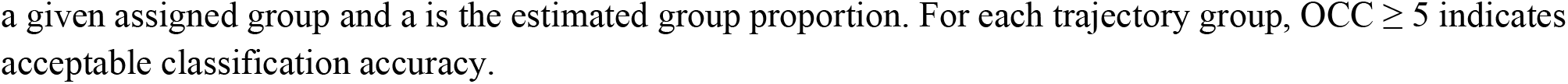
Fit statistics for group-based multi-trajectory models of ALT, albumin, and total bilirubin.

| Trajectory groups | BIC | Sample size-adjusted BIC | AIC | Group membership % | Group AvePP b | OCC c |
| --- | --- | --- | --- | --- | --- | --- |
| 2 | 5265.148 | 5198.495 | 5177.238 | 69.75 | 0.979–0.995 | 21.09–433.58 |
| 2 | 5265.148 | 5198.495 | 5177.238 | 30.25 | 0.979–0.995 | 21.09–433.58 |
| 3 | 4774.793 | 4660.531 | 4624.090 | 15.43 | 0.972–0.996 | 41.36–419.43 |
| 3 | 4774.793 | 4660.531 | 4624.090 | 46.91 | 0.972–0.996 | 41.36–419.43 |
| 3 | 4774.793 | 4660.531 | 4624.090 | 37.65 | 0.972–0.996 | 41.36–419.43 |
| 4 | 4682.009 | 4526.485 | 4476.885 | 15.43 | 0.962–0.990 | 59.58–532.51 |
| 4 | 4682.009 | 4526.485 | 4476.885 | 38.89 | 0.962–0.990 | 59.58–532.51 |
| 4 | 4682.009 | 4526.485 | 4476.885 | 15.43 | 0.962–0.990 | 59.58–532.51 |
| 4 | 4682.009 | 4526.485 | 4476.885 | 30.25 | 0.962–0.990 | 59.58–532.51 |
| 5 | 4725.385 | 4519.079 | 4453.282 | 14.81 | 0.967–1.000 | 45.95–582.65 |
| 5 | 4725.385 | 4519.079 | 4453.282 | 39.51 | 0.967–1.000 | 45.95–582.65 |
| 5 | 4725.385 | 4519.079 | 4453.282 | 14.20 | 0.967–1.000 | 45.95–582.65 |
| 5 | 4725.385 | 4519.079 | 4453.282 | 1.85 | 0.967–1.000 | 45.95–582.65 |
| 5 | 4725.385 | 4519.079 | 4453.282 | 29.63 | 0.967–1.000 | 45.95–582.65 |

\* Notes: BIC = Bayesian information criterion; AIC = Akaike information criterion; AvePP = average posterior probability; OCC = odds of correct classification. a For all models, the polynomial order for all trajectory classes was set to quadratic. b Group AvePP of assignment: Based on the maximum posterior probability assignment rule, each individual was assigned to the trajectory group for which the posterior probability was the largest. For each trajectory group, an AvePP > 0.70 indicates good classification accuracy. c OCC was calculated as: $OCC = (b/(1-b))/(a/(1-a))$ where b is the average posterior probability for
a given assigned group and $a$ is the estimated group proportion. For each trajectory group, $OCC \geq 5$ indicates acceptable classification accuracy.

### Characteristics of Trajectory Patterns

The four trajectory classes showed distinct longitudinal patterns. Overall, between-group separation was primarily driven by differences in ALT and total bilirubin, while albumin remained relatively stable across all groups. Class 1 was characterized by relatively stable ALT and bilirubin levels in the early phase, followed by an increase at later time points. Class 2 showed a decreasing ALT trend with a subsequent increase in bilirubin. Class 3 demonstrated a transient decrease in ALT with partial recovery, while bilirubin remained relatively stable. Class 4 exhibited relatively stable ALT but progressively increasing bilirubin over time.

**Table 3.** Description of the multi-trajectory classes based on ALT, albumin, and total bilirubin.

| Trajectory group | N (%) | Biomarker | Description and standardized pattern from time point 1 to time point 5 |
| --- | --- | --- | --- |
| Class 1 | 38.9 | ALT | Near baseline at time point 1, slightly below baseline at time point 3, and then markedly increased by time point 5, indicating a late-rising ALT pattern. |
| Class 1 | 38.9 | Albumin | Mildly above baseline and essentially stable across all time points, suggesting a preserved short-term albumin trajectory. |
| Class 1 | 38.9 | Total bilirubin | Slightly below baseline at time points 1 and 3, followed by a sharp rise by time point 5, consistent with delayed bilirubin elevation. |
| Class 2 | 15.4 | ALT | Near baseline at time point 1, then progressively decreased over time and reached the lowest level at time point 5, representing a pronounced downward ALT trajectory. |
| Class 2 | 15.4 | Albumin | Mildly above baseline and stable throughout follow-up, with minimal temporal variation. |
| Class 2 | 15.4 | Total bilirubin | Slightly below baseline at time points 1 and 3, followed by a steep increase at time point 5, indicating substantial late bilirubin worsening. |
| Class 3 | 15.4 | ALT | Near baseline at time point 1, sharply decreased at time point 3, and partially recovered by time point 5, although it remained below baseline overall. |
| Class 3 | 15.4 | Albumin | Mildly above baseline and stable across all time points. |
| Class 3 | 15.4 | Total bilirubin | Slightly below baseline at time point 1 and remained persistently low with minimal fluctuation thereafter, suggesting a stable low-bilirubin trajectory. |
| Class 4 | 30.2 | ALT | Close to baseline at time point 1 and remained essentially flat thereafter, indicating a stable ALT trajectory. |
| Class 4 | 30.2 | Albumin | Mildly above baseline and stable throughout follow-up. |
| Class 4 | 30.2 | Total bilirubin | Slightly below baseline at time point 1, crossed above baseline by time point 3, and continued to rise by time point 5, representing a steadily increasing bilirubin trajectory. |

**Figure 1.**
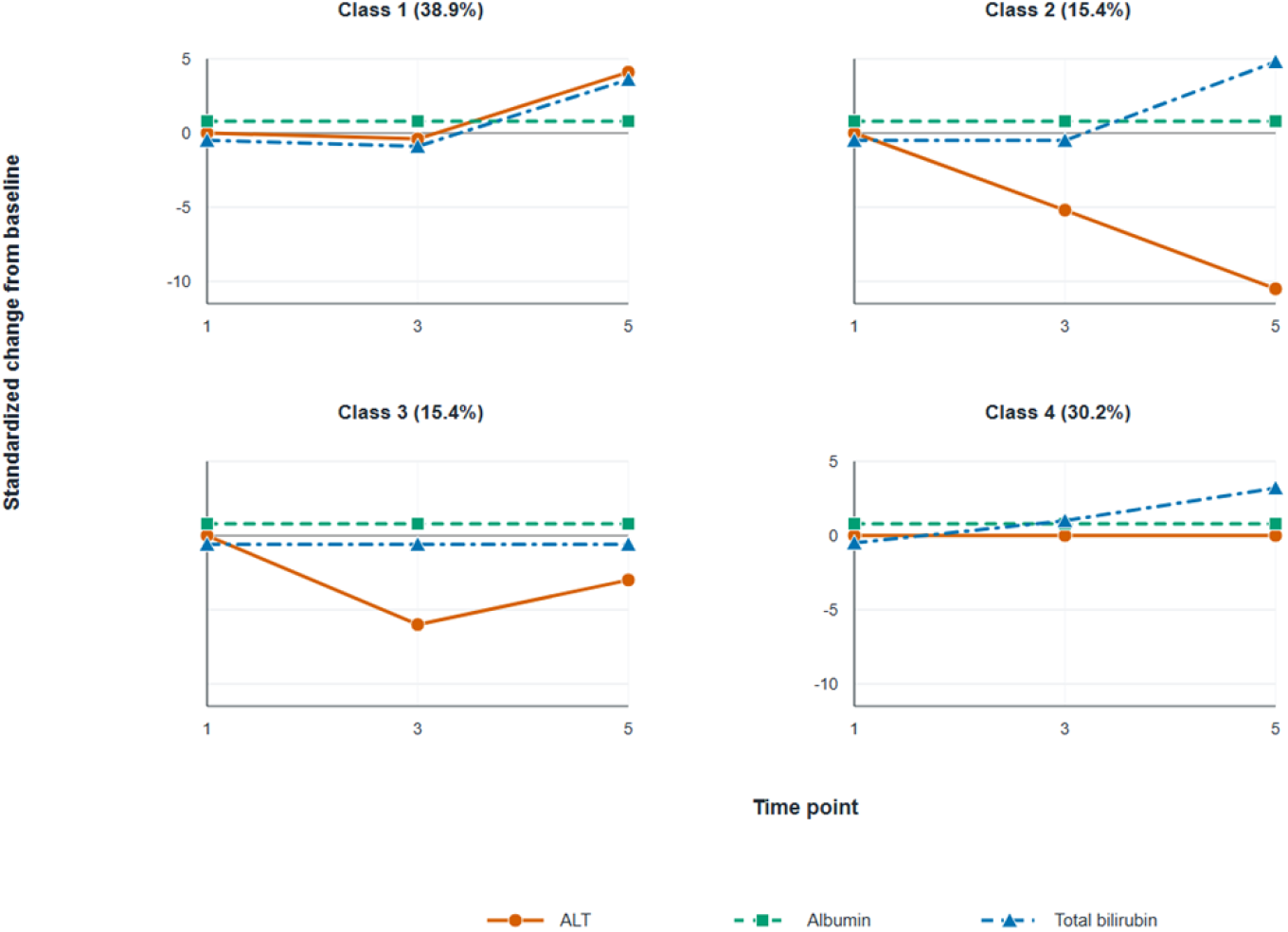
Trajectory patterns of ALT, albumin, and total bilirubin across the four latent classes, shown as standardized change from baseline. Between-class separation was mainly driven by dynamic changes in ALT and total bilirubin, whereas albumin showed limited short-term variation.

### Time to Diagnosis of SALI Across Trajectory Classes

The time from ICU admission to the first diagnosis of SALI differed significantly across trajectory classes. Class 2 had the shortest time to diagnosis, whereas Class 1 had the longest. Class 3 and Class 4 showed intermediate values.

**Figure 2.**
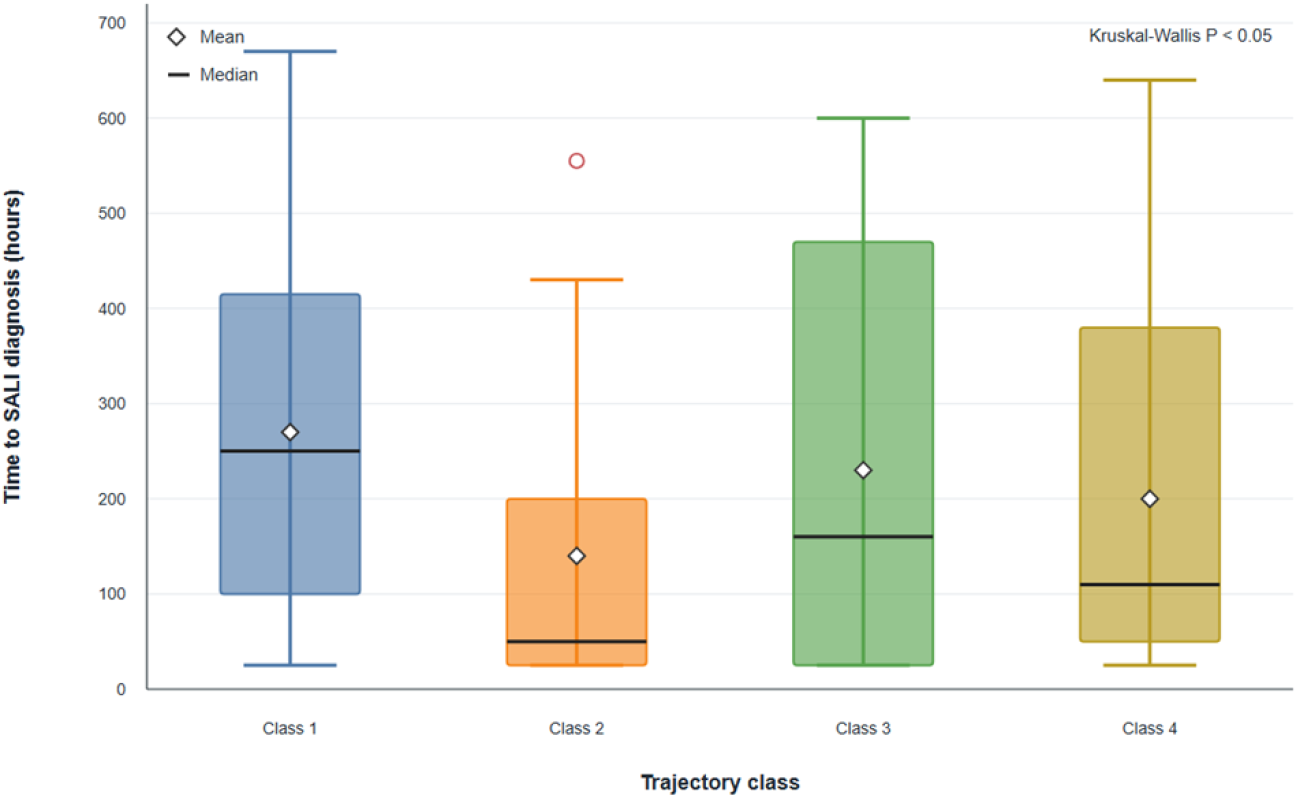
Time to diagnosis of sepsis-associated liver injury (SALI) across trajectory classes. Boxplots show the distribution of time (hours) from ICU admission to SALI diagnosis in each trajectory group. The median is indicated by the horizontal line, and boxes represent the interquartile range (IQR), with whiskers extending to 1.5 × IQR. Outliers are displayed as individual points. A significant difference was observed among groups (P < 0.05).

### Baseline Characteristics Across Trajectory Classes

Among the laboratory parameters on day 1 across the different multi-trajectory classifications, ALT levels were lowest in Class 1 patients and highest in Class 2 patients (P < 0.001). Total bilirubin was significantly elevated in Class 3 patients and lowest in Class 2 patients (P < 0.001). Albumin levels were lowest in Class 1 patients and relatively higher in Class 3 patients (P < 0.001). Regarding coagulation and liver function parameters, there were significant differences in INR and prothrombin time (PT) across groups (all P < 0.001), with Class 2 patients having the highest INR values. Both direct and indirect bilirubin levels were significantly elevated in Class 3 patients (both P < 0.001). In contrast, no significant differences were observed in AST or lactate dehydrogenase (LDH) levels. Regarding organ function, there were significant differences in lactate levels across groups (P = 0.024), with the highest levels observed in Class 4 patients.The proportion of patients receiving mechanical ventilation was highest in Class 4 (95.65%), although this difference did not reach statistical significance. Regarding demographic characteristics and comorbidities, there was a significant difference in age among the groups (P < 0.001), with patients in Class 3 being younger. The prevalence of heart failure also differed significantly, with the highest proportion observed in Class 2 (P < 0.001). There were no significant differences among the groups in terms of gender distribution or severity scores (SAPS II).

**Table 4.** Baseline characteristics across trajectory classes.

| Variable | Class 1 (n=63) | Class 2 (n=25) | Class 3 (n=25) | Class 4 (n=49) | P value |
| --- | --- | --- | --- | --- | --- |
| ALT | 1.40(1.25 - 1.50) | 2.45<br>(1.90 - 3.30) | 2.10<br>(1.70 - 2.55) | 2.00<br>(1.70 - 2.85) | <0.001 |
| Total bilirubin | 1.90(0.70-3.85) | 1.50(0.90 - 1.85) | 24.65(19.40- 31.00) | 6.60 (4.10 - 9.40) | <0.001 |
| Albumin, | 2.45(2.20- 2.90) | 2.95(2.45 - 3.50) | 3.10(2.80 - 3.80) | 3.00 (2.50 - 3.50) | <0.001 |
| INR | 1.40(1.20 - 1.50) | 2.30(1.60 - 3.40) | 1.90(1.70 - 2.40) | 2.00 (1.60 - 2.50) | <0.001 |
| PTT | 35.10(29.90-43.80) | 39.30(34.00 - 46.70) | 40.00(36.10 - 47.30) | 39.35(34.90 - 50.40) | 0.109 |
| PT | 14.80(13.50- 16.90) | 23.80(18.00 - 36.70) | 20.70(18.40 - 26.30) | 21.60(18.00 - 26.70) | <0.001 |
| Direct bilirubin | 2.10(1.20 - 4.20) | 1.10(1.00 - 2.00) | 20.40(13.00 - 24.60) | 4.60 (2.85 - 6.75) | <0.001 |
| Indirect bilirubin | 0.70(0.55 - 1.00) | 1.10(0.60 - 1.20) | 8.75(6.45 - 10.35) | 2.40 (1.40 - 3.85) | <0.001 |
| Alkaline phosphatase | 76.00(57.00- 117.00) | 115.00(73.00 - 154.00) | 113.00(91.00 - 227.00) | 109.50(70.00 - 171.00) | 0.045 |
| AST | 91.00(45.00 - 265.00) | 58.50(34.50 - 770.00) | 101.00(70.00 - 194.00) | 133.50(59.00 - 459.00) | 0.579 |
| LDH | 418.50 (259.00 - 726.00) | 496.00 (198.00 - 1,850.00) | 380.00(227.00 - 491.00) | 354.00 (258.00 - 795.00) | 0.366 |
| Platelet count | 157.00 (101.00 - 220.00) | 166.00 (110.00 - 255.00) | 136.00(61.00 - 203.00) | 137.50 (87.00 - 181.00) | 0.081 |
| White blood cell | 11.00(6.30 - 16.10) | 13.80(8.30 - 16.40) | 14.10(8.90 - 22.50) | 10.95(6.70- 14.70) | 0.264 |
| Hemoglobin | 9.40(8.50 - 11.80) | 10.40(8.50 - 11.40) | 9.20(8.30 - 10.10) | 8.50(7.95- 10.15) | 0.200 |
| Lactate | 1.90(1.40 - 2.90) | 2.65(1.85 - 4.45) | 2.50(1.70 - 3.40) | 2.80 (1.80 - 6.85) | 0.024 |
| C-reactive protein | 165.80(119.40 - 253.50) | 40.20(40.20 - 40.20) | 76.50(58.50 - 94.50) | 107.00(97.60- 140.80) | 0.070 |
| Creatinine | 1.40(0.90 - 3.80) | 1.60(1.20 - 3.50) | 2.00(1.10 - 4.30) | 1.35 (0.90 - 2.00) | 0.285 |
| Acute kidney | 51.00 (87.93%) | 19.00 (90.48%) | 21.00 (91.30%) | 45.00 (97.83%) | 0.326 |
| CRRT, n (%) | 19.00 (32.76%) | 8.00 (38.10%) | 6.00 (26.09%) | 16.00 (34.78%) | 0.847 |
| Mechanical ventilation, n (%) | 47.00 (81.03%) | 18.00 (85.71%) | 20.00 (86.96%) | 44.00 (95.65%) | 0.176 |
| Age | 62.28(52.27 - 72.79) | 63.90(58.95 - 70.03) | 46.89(40.54 - 53.88) | 54.95(48.20- 62.30) | <0.001 |
| Sex, n (%) |  |  |  |  | 0.820 |
| Female | 20.00 (34.48%) | 9.00 (42.86%) | 10.00 (43.48%) | 19.00 (41.30%) |  |
| Male | 38.00 (65.52%) | 12.00 (57.14%) | 13.00 (56.52%) | 27.00 (58.70%) |  |
| Congestive heart failure, n (%) | 21.00 (36.21%) | 12.00 (57.14%) | 1.00 (4.35%) | 9.00 (19.57%) | <0.001 |
| SAPS II, | 43.00(35.00- 50.00) | 44.00(34.00- 63.00) | 49.00(31.00- 55.00) | 45.00(35.00- 65.00) | 0.857 |

Continuous variables are presented as median (interquartile range), and categorical variables are presented as counts (percentages). Comparisons between groups were performed using the Kruskal-Wallis test for continuous variables and the chi-square test or Fisher’s exact test for categorical variables, as appropriate.

Patients with fewer than three repeated measurements of ALT, total bilirubin, or albumin during the first 7 days were excluded before trajectory modeling. For regression analyses, variables with available data were used in complete-case models. Kaplan-Meier survival curves and landmark analysis were used to compare 60-day survival among trajectory classes [27]. The day-22 landmark was selected as an exploratory time point based on the observed separation of survival curves and was used to reduce time-related bias when comparing post-landmark mortality. Cox proportional hazards regression models were used to evaluate the association between trajectory class membership and 60-day mortality [25]. Multivariable models were adjusted for clinically relevant covariates selected a priori based on clinical experience, including age, lactate, renal disease, and SOFA score. The proportional hazards assumption was assessed using Schoenfeld residuals and weighted residual-based diagnostics [26,28]. A two-sided P value <0.05 was considered statistically significant. All analyses were performed using R software (version 4.5.1).

### Landmark Analysis

Before the day-22 landmark, survival did not differ significantly across the four trajectory classes. After day 22, Class 3 and Class 4 showed substantially lower survival than Class 1. In Cox regression, Class 3 (HR 4.374, 95% CI 1.960-9.759, P < 0.001) and Class 4 (HR 7.451, 95% CI 3.649-15.212, P < 0.001) were associated with increased mortality risk after the landmark time point, whereas Class 2 was not significantly different from Class 1.

**Figure 3.**
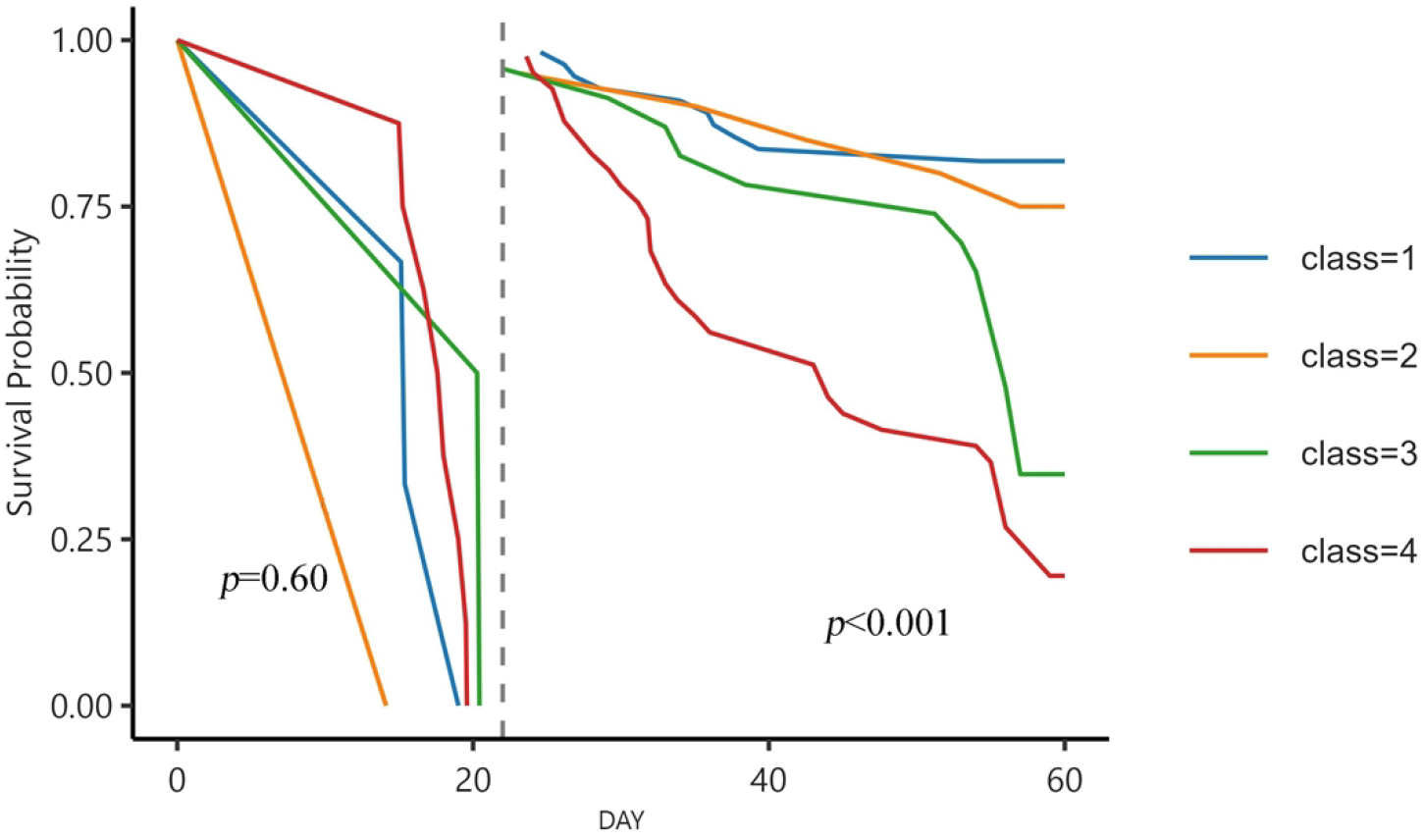
Kaplan-Meier-like fitted survival curves for the four multi-trajectory classes. The vertical dashed line at day 22 denotes the landmark time. Class 1 (reference) shows consistently high survival; Classes 3 and 4 diverge downward after day 22, with Class 4 showing the lowest survival. Class 2 remains close to Class 1.

**Table 5.** Association between multi-trajectory class and mortality risk before and after the landmark time point.

| Variable | Before Landmark Time | Before Landmark Time | After Landmark Time | After Landmark Time |
| --- | --- | --- | --- | --- |
| Variable | HR (95% CI) | P value | HR (95% CI) | P value |

| Multi-trajectory group |  |  |  |  |
| --- | --- | --- | --- | --- |
| Class 1 | Reference |  | Reference |  |
| Class 2 | 0.934 (0.488–3.381) | 0.953 | 1.391 (0.475–4.07) | 0.541 |
| Class 3 | 1.504 (1.777–9.004) | 0.655 | 4.374 (1.960–9.759) | <0.001 |
| Class 4 | 3.235(0.882–12.534) | 0.676 | 7.451(3.649–15.212) | <0.001 |

\*. HR, hazard ratio; CI, confidence interval. The multi-trajectory classes were derived from longitudinal patterns of ALT, albumin, and total bilirubin. Class 1 served as the reference.

## Discussion

In this study, we identified four distinct joint trajectory patterns of alanine aminotransferase (ALT), total bilirubin, and albumin in patients with sepsis-associated liver injury and demonstrated that these patterns were associated with significant differences in patient mortality after day 22. In particular, trajectory patterns characterized by persistently elevated or progressively increasing total bilirubin were associated with a substantially higher risk of death.

Previous studies have primarily evaluated liver dysfunction in sepsis using static measurements of biomarkers, such as total bilirubin or transaminases ^[5-6,9-11,29-31]^. While these studies have consistently shown that abnormal liver function is associated with poor outcomes, they do not capture the temporal evolution of organ dysfunction ^[5,8,32-34]^. In contrast, trajectory-based approaches incorporate longitudinal information and have been shown to improve prognostic assessment in critically ill patients, particularly in studies of lactate and SOFA score trajectories ^[16-18,31]^. Our findings extend this framework to liver-related biomarkers and suggest that incorporating temporal patterns may provide additional insight beyond single time-point assessments ^[16,33]^.

This study found that, among laboratory indicators on the first day of admission, when grouped into survival and mortality cohorts, the vast majority of baseline indicators showed no significant differences between the two groups, including liver function indicators, coagulation parameters, renal function, and organ support treatment modalities. Albumin was the only liver-related biomarker showing a significant difference between the two groups, with albumin levels in the mortality group being slightly higher than those in the survival group. A possible reason for this is that the mortality group was predominantly characterized by cholestatic liver injury, with relatively preserved early synthetic function. When patients were grouped into multiple trajectories based on ALT, albumin, and total bilirubin, one group had the lowest albumin levels among the different trajectory classifications, while Class 3 had relatively higher albumin levels. In the Kaplan-Meier analysis, Class 1 had a higher mortality rate than Class 3. This finding is consistent with previous studies, indicating that a single baseline albumin measurement has limited prognostic value and highlighting the necessity of dynamic trajectory analysis using multiple indicators.

This study constructed a combined trajectory model based on ALT, albumin, and total bilirubin and found that differences in trajectory grouping were primarily driven by ALT and total bilirubin, while early fluctuations in albumin were relatively small. This characteristic is consistent with the physiological functions and pathological response patterns of the three markers. Total bilirubin reflects hepatic excretory function and tends to show persistent changes during the course of the disease; ALT represents the level of hepatocyte injury and fluctuates significantly with changes in inflammation and disease severity; whereas albumin has a long half-life, is minimally affected by acute pathological states in the short term, and exhibits relatively stable dynamic changes. Baseline values at admission showed significant heterogeneity across trajectory subgroups: patients in Class 1 had the lowest ALT and albumin levels, suggesting mild hepatocyte injury but weak hepatic synthetic reserve and poor baseline nutritional status; Class 2 patients had the highest ALT levels and low total bilirubin, along with elevated INR and a higher prevalence of heart failure; their primary phenotype was acute hepatocyte injury, often complicated by coagulation disorders and cardiac dysfunction; Class 3 patients were younger, had significantly elevated total bilirubin, and higher baseline albumin levels, presenting a phenotype characterized by cholestasis with relatively intact early hepatic synthetic function; Class 4 patients had the highest lactate levels, with more severe systemic tissue hypoperfusion and a more severe sepsis-induced stress response ^[6-8,11,35]^.

Landmark analysis and multivariable regression results showed that the prognosis for Class 4 patients was worst after day 22; prior to day 22, there were no significant differences between the four groups; compared with Class 1, patients in Groups 3 and 4 had a significantly higher risk of death. In summary, subtypes based on dynamic liver function trajectories driven by ALT and total bilirubin can effectively distinguish the clinical phenotypes and severity of sepsis-associated liver injury, and provide valuable early warning and risk stratification for mortality outcomes 20 days after hospital admission ^[25-28]^.

The time to diagnosis of sepsis-associated liver injury varied across different trajectory classes. Class 2 had the earliest time to diagnosis, which was closely related to its unique clinical phenotype. Class 2 was characterized by the highest ALT levels and the lowest total bilirubin levels, presenting a liver injury pattern dominated by acute hepatocellular injury. As an early and sensitive marker of hepatocyte injury, ALT can rise rapidly under sepsis-induced stress, making it easier to meet the diagnostic criteria for sepsis-associated liver injury at an early stage. Concurrently, this group exhibited a marked elevation in INR and a higher prevalence of heart failure; the combination of coagulation disorders and cardiac dysfunction further exacerbated acute stress-induced liver injury, accelerating the clinical manifestation of the liver injury phenotype. Compared to other subgroups, Class 2 presented with acute hepatocyte injury as the initial manifestation, with laboratory abnormalities appearing earlier and a lower clinical identification threshold, making it the subtype with the earliest time to definitive diagnosis of sepsis-associated liver injury. However, this was not consistently associated with mortality risk. This suggests that the dynamic trajectory of biomarker changes can provide prognostic information beyond the mere time to diagnosis21.

This study utilized repeated measurements to describe temporal changes in liver function, thereby providing a more comprehensive view of disease progression; secondly, the combined trajectory modeling approach allowed for the simultaneous evaluation of multiple biomarkers, capturing different dimensions of liver dysfunction; thirdly, this study linked these dynamic patterns to clinically relevant outcomes and employed threshold-based survival analysis to eliminate unmeasured error from time-dependent variables. At the same time, several limitations of this study should be acknowledged. This is a single-center retrospective study, which may be subject to selection bias and information bias, and does not allow for causal inferences. The sample size is relatively small, with a limited number of patients in some trajectory classes, which may affect the stability of the estimates. Furthermore, not all relevant liver biomarkers were included, and no external validation was performed. Therefore, the generalizability of the study results remains uncertain.

## Conclusions

In summary, this study identified distinct combined clinical course patterns of ALT, total bilirubin, and albumin in patients with sepsis-associated liver injury, highlighting the dynamic heterogeneity of liver dysfunction in this population. Clinical course patterns characterized by persistently elevated or gradually rising bilirubin levels were associated with higher mortality rates after day 22.

## Data Availability

No datasets were generated or analysed during the current study. All relevant data from this study will be made available upon study completion.

## Data Availability

Due to institutional and privacy restrictions involving anonymized clinical records, the underlying data are available from the corresponding author upon reasonable request and with appropriate institutional approval.

## Funding

The authors received no specific funding for this work.

## Competing Interests

The authors have declared that no competing interests exist.

## Author Contributions

Conceptualization: Yanan Qi, Cheng Liu. Data curation: Yanan Qi, Tian Zhou, Qinmin Zhao. Formal analysis: Yanan Qi. Methodology: Yanan Qi, Cheng Liu. Supervision: Cheng Liu. Writing - original draft: Yanan Qi. Writing - review and editing: Tian Zhou, Qinmin Zhao, Cheng Liu.

## Acknowledgments

None.

## Notes

### Competing Interest Statement

The authors have declared no competing interest.

### Clinical Trial

N

### Funding Statement

The author(s) received no specific funding for this work.

### Author Declarations

he studies involving human participants were reviewed and approved by the Medical Ethics Committee of The First Affiliated Hospital of Bengbu Medical University (Approval No. [2023]342). The study was conducted in accordance with the principles of the Declaration of Helsinki. Written informed consent was not required because this was a retrospective study using anonymized clinical data collected during routine care. The data were fully anonymized prior to the researchers’ access, and no direct patient identifiers were obtained. The study did not involve direct patient contact, intervention, or collection of additional biological samples. The requirement for written informed consent was waived by the Medical Ethics Committee of The First Affiliated Hospital of Bengbu Medical University in accordance with national legislation and institutional requirements.

## References

[1] Singer M, Deutschman CS, Seymour CW, et al. The Third International Consensus Definitions for Sepsis and Septic Shock (Sepsis-3). JAMA. 2016;315(8):801–810. doi:10.1001/jama.2016.0287.

[2] Srzic I, Nesek Adam V, Tunjic Pejak D. Sepsis definition: what is new in the treatment guidelines. Acta Clin Croat. 2022;61(Suppl 1):67–72. doi:10.20471/acc.2022.61.s1.11.

[3] Evans L, Rhodes A, Alhazzani W, et al. Surviving Sepsis Campaign: international guidelines for management of sepsis and septic shock 2021. Intensive Care Med. 2021;47(11):1181–1247. doi:10.1007/s00134-021-06506-y.

[4] Lei J, Xia X, Cheng X, Qu H, Chen Z. Supervised machine learning models for predicting sepsis-associated liver injury in patients with sepsis: development and validation study based on a multicenter cohort study. J Med Internet Res. 2025;27:e66733. doi:10.2196/66733.

[5] Kramer L, Jordan B, Druml W, Bauer P, Metnitz PGH. Incidence and prognosis of early hepatic dysfunction in critically ill patients: a prospective multicenter study. Crit Care Med. 2007;35(4):1099–1104. doi:10.1097/01.CCM.0000259462.97164.A0.

[6] Nesseler N, Launey Y, Aninat C, Morel F, Mallédant Y, Seguin P. Clinical review: the liver in sepsis. Crit Care. 2012;16(5):235. doi:10.1186/cc11381.

[7] Recknagel P, Gonnert FA, Westermann M, et al. The liver in sepsis: shedding light on the cellular basis of hepatocyte dysfunction. Crit Care. 2013;17(4):153. doi:10.1186/cc12731.

[8] Horvatits T, Drolz A, Trauner M, Fuhrmann V. Liver injury and failure in critical illness. Hepatology. 2019;70(6):2204–2215. doi:10.1002/hep.30824.

[9] Patel JJ, Taneja A, Niccum D, Kumar G, Jacobs E, Nanchal R. The association of serum bilirubin levels on the outcomes of severe sepsis. J Intensive Care Med. 2015;30(1):23–29. doi:10.1177/0885066613488739.

[10] Yang ZX, Lv XL, Yan J. Serum total bilirubin level is associated with hospital mortality rate in adult critically ill patients: a retrospective study. Front Med (Lausanne). 2021;8:697027. doi:10.3389/fmed.2021.697027.

[11] Chand N, Sanyal AJ. Pathophysiology of sepsis-induced cholestasis: a review. JGH Open. 2022;6(6):378–387. doi:10.1002/jgh3.12771.

[12] Zhang X, Wang T, Gui P, et al. The gut-liver axis in sepsis: interaction mechanisms and therapeutic potential. Crit Care. 2022;26(1):213. doi:10.1186/s13054-022-04090-1.

[13] Lee HJ, Jang Y, Jung H, et al. Modified cardiovascular SOFA score in sepsis: development and internal and external validation. BMC Med. 2022;20(1):263. doi:10.1186/s12916-022-02461-7.

[14] Wen C, Zhang A, Hu B, et al. An interpretable machine learning model for predicting 28-day mortality in patients with sepsis-associated liver injury. PLoS One. 2024;19(5):e0303469. doi:10.1371/journal.pone.0303469.

[15] Nagin DS, Jones BL, Passos VL, Tremblay RE. Recent advances in group-based trajectory modeling for clinical research. Annu Rev Clin Psychol. 2024;20:285–305. doi:10.1146/annurev-clinpsy-081122-012416.

[16] Nagin DS, Jones BL, Passos VL, Tremblay RE. Group-based multi-trajectory modeling. Stat Methods Med Res. 2018;27(7):2015–2023. doi:10.1177/0962280216673085.

[17] Magrini A. Assessment of agricultural sustainability in European Union countries: a group-based multivariate trajectory approach. Adv Stat Anal. 2022;106:673–703. doi:10.1007/s10182-022-00437-9.

[18] Magrini A. gbmt: Group-Based Multivariate Trajectory Modeling. R package version 0.1.4. 2024. Available from: https://CRAN.R-project.org/package=gbmt.

[19] Na AY, Yoon SH, Lee YJ, et al. Novel time-dependent multi-omics integration in sepsis-associated liver dysfunction. Genomics Proteomics Bioinformatics. 2023;21(6):1101–1116. doi:10.1016/j.gpb.2023.04.002.

[20] Gou E, Liu Y, Zhang Y, Wang X, Li J. Association between albumin-bilirubin score and in-hospital mortality in patients with sepsis: evidence from two large databases. Heliyon. 2024;10(15):e34697. doi:10.1016/j.heliyon.2024.e34697.

[21] Xiao Y, Li W, Zhang Y, Chen Y, Zhang L. Evaluation of qSOFA score, conjugated bilirubin, and creatinine levels for predicting 28-day mortality in patients with sepsis. Exp Ther Med. 2022;24(1):447. doi:10.3892/etm.2022.11374.

[22] Wang J, Zhang H, Chen X, Li Y, Zhao Y. Predictive role of the albumin-bilirubin score in ICU patients with cirrhosis and sepsis: insights from a large retrospective cohort. BMC Gastroenterol. 2025;25(1):520. doi:10.1186/s12876-025-04111-7.

[23] Palmowski L, Schuttler J, Lehmann F, et al. Subphenotypes and the De Ritis ratio for mortality risk stratification in sepsis-associated acute liver injury: a retrospective cohort study. EClinicalMedicine. 2025;82:103173. doi:10.1016/j.eclinm.2025.103173.

[24] Zhang Z, Zhang X, Luo Y, Chen Y. Bibliometric study of sepsis-associated liver injury from 2000 to 2023. World J Gastroenterol. 2024;30(30):3609–3624. doi:10.3748/wjg.v30.i30.3609.

[25] Cox DR. Regression models and life-tables. J R Stat Soc Series B Stat Methodol. 1972;34(2):187–202. doi:10.1111/j.2517-6161.1972.tb00899.x.

[26] Schoenfeld D. Partial residuals for the proportional hazards regression model. Biometrika. 1982;69(1):239–241. doi:10.1093/biomet/69.1.239.

[27] van Houwelingen HC. Dynamic prediction by landmarking in event history analysis. Scand J Stat. 2007;34(1):70–85. doi:10.1111/j.1467-9469.2006.00529.x.

[28] Grambsch PM, Therneau TM. Proportional hazards tests and diagnostics based on weighted residuals. Biometrika. 1994;81(3):515–526. doi:10.1093/biomet/81.3.515.

[29] Geier A, Fickert P, Trauner M. Mechanisms of disease: mechanisms and clinical implications of cholestasis in sepsis. Nat Clin Pract Gastroenterol Hepatol. 2006;3(10):574–585. doi:10.1038/ncpgasthep0602.

[30] Choi S, Kim WY, Ryoo SM, et al. Prognostic value of the AST/ALT ratio in patients with septic shock: a prospective, multicenter, registry-based observational study. Diagnostics. 2025;15(14):1773. doi:10.3390/diagnostics15141773.

[31] Elmi AN, Kwo PY. The liver in sepsis. Clin Liver Dis. 2025;29(3):453–467. doi:10.1016/j.cld.2025.04.002.

[32] Li X, Zhang Y, Wang H, Liu J. Clinical characteristics and prognostic factors of sepsis-associated liver injury. Zhonghua Wei Zhong Bing Ji Jiu Yi Xue. 2024;36(11):1121–1126. doi:10.3760/cma.j.cn121430-20240426-00386.

[33] Perez Ruiz de Garibay A, Drolz A, Wendon J, Karvellas CJ. Critical care hepatology: definitions, incidence, prognosis and role of liver failure in critically ill patients. Crit Care. 2022;26(1):289. doi:10.1186/s13054-022-04163-1.

[34] Beyer D, Hoff J, Sommerfeld O, Zipprich A, Gassler N, Press AT. The liver in sepsis: molecular mechanism of liver failure and their potential for clinical translation. Mol Med. 2022;28(1):84. doi:10.1186/s10020-022-00510-8.

[35] Papathanakos G, Andrianopoulos I, Papathanasiou A, Koulouras V. Clinical sepsis phenotypes in critically ill patients. Microorganisms. 2023;11(9):2165. doi:10.3390/microorganisms11092165.

